# Comprehensive profiling of the mutational landscape of hidradenoma papilliferum validates key role of alterations in the PI3K/AKT pathway alterations

**DOI:** 10.1101/2025.03.31.25324887

**Authors:** Saamin Cheema, Louise van der Weyden, Kim Wong, Martin Del Castillo Velasco-Herrera, Jamie Billington, Ian Vermes, Elizabeth Anderson, Laura Allen, Nicolas de Saint Aubain, Michiel P. J. van der Horst, Ahmed K. Alomari, Carolin Mogler, Carlos Monteagudo, Derek Frew, Steven D. Billings, Mark J. Arends, Ingrid Ferreira, Thomas Brenn, David J. Adams

## Abstract

**Background:** Hidradenoma papilliferum (HP) is a rare, benign epithelial tumour arising from the anogenital mammary-like glands, that occurs almost exclusively in the anogenital region of middle-aged women. To-date, genetic characterisation of HP has identified recurrent somatic mutations in *PIK3CA* and *AKT1,* which appear to be mutually exclusive, and *PIK3R1* mutations in some cases without *PIK3CA* or *AKT1* mutations. However, these studies used small sample sizes and/or analysed a limited number of genes, thus the mutational landscape of HP remains incomplete.

**Objectives:** To better understand the molecular pathogenesis of HP by exploring the mutational landscape of these rare tumours and identifying candidate driver events.

**Methods:** In this retrospective, multi-institutional study, we collated a cohort of formalin-fixed paraffin-embedded HP cases (n=68) to perform whole-exome sequencing and RNA-sequencing.

**Results:** *PIK3CA* and *PIK3R1* were identified as significantly mutated, and thus, are statistically predicted driver genes of HP. The mutations, which were predominantly at known hotspots, were all predicted to be oncogenic/likely oncogenic. Mutation of *PIK3CA* and *PIK3R1* were mutually exclusive, consistent with these genes each encoding a subunit of the heterodimeric enzyme, PI3 kinase (PI3K), which functions via the PI3K/AKT/mTOR signalling pathway. Somatic mutations were also identified in other members of the PI3K/AKT/mTOR pathway (*AKT1/2*), receptors or receptor-associated proteins that signal via the PI3K/ATK/mTOR pathway (*ERBB3, IRS2*), or members of pathways that crosstalk with the PI3K/AKT/mTOR pathway (*AR* and *MAPK1*). The copy number landscape of HP revealed no recurrent alterations. Fusion gene analysis identified fusions in individual samples (*SMARCD3::ASXL2, STX17::ERO1B, ANKFY1::UBE2G1, IQGAP1::ZNF774* and *EXOC2::FRMD4B*), with only the latter two being previously reported in other cancer types. No significant evidence of human papillomavirus (HPV) sequence was found in the tumour samples.

**Conclusions:** Comprehensive characterisation of the mutational landscape of HP identified *PIK3CA* and *PIK3R1* as mutually exclusive driver genes. Taken together with mutation of other genes involved in PI3K/AKT signalling, our findings confirm a key role of the alteration of this pathway in the pathogenesis of HP. The absence of HPV sequences suggests HPV is not involved in the aetiology of HP.

## INTRODUCTION

Hidradenoma papilliferum (HP) is a rare, benign epithelial tumour arising from the anogenital mammary-like glands (AGMLG). It occurs almost exclusively in the anogenital region, predominantly on the vulva, of middle-aged women.^1,2^ There have only been a few isolated reports of anogenital HP in male patients.^3,4^ HP most commonly presents as a solitary, asymptomatic, slow-growing, well-circumscribed, skin-coloured to red nodule, rarely exceeding 2 cm in diameter.^1,2,5^ HP is difficult to diagnose solely on clinical presentation as it mimics many other cutaneous neoplasms, and as such, histopathological examination of the lesion is an essential part of the diagnosis.^1,6^ Management of HP is local excision and carries a favourable prognosis, however, incomplete excision has been associated with local recurrence.^7^ Malignant transformation of HP is rare, limited to a few case reports.^8–10^

The aetiology of HP is currently unclear. Human papillomavirus (HPV) DNA has been detected in some anogenital HP lesions^11,12^ but not in others,^13–15^ suggesting it does not play a causative role. Some studies have shown that HP are oestrogen receptor (ER) and progesterone receptor (PR) immunopositive,^16,17^ although other studies have reported ER and/or PR immunonegativity,^18,19^ thus a possible relationship between HP and hormone receptors is unclear.

To-date there have been five studies investigating the genetics of HP. Two studies have performed Sanger sequencing of 7-30 cases to look at the mutation status of four genes (*PIK3CA*, *AKT1*, *RAS*, and *BRAF*)^20,21^ and three studies have performed next generation sequencing (NGS) of 5-15 cases using targeted panels of 50-409 cancer-associated genes or specific exons encompassing known mutation hotspots.^14,15,22^ These studies have found that *PIK3CA* and *AKT1* are frequently mutated in HP (ranging from 20-63% and 3-14% cases, respectively), and these mutations appear to be mutually exclusive.^14,15,20–22^ Additionally, investigation of *PIK3CA/AKT1* mutation negative HP cases found mutations in *PIK3R1* (3/7 cases) and other genes (1/7 cases each) with putative links to the PI3K/AKT signalling pathway, proposing this pathway as a key mediator of HP tumourigenesis.^14^

However, Sanger sequencing and NGS using targeted panels only offers us a partial view of the underlying genetics of these tumours. Thus, to comprehensively characterise the mutational landscape of HP, we collated a cohort of 68 formalin-fixed paraffin-embedded (FFPE) HP cases, from which samples were taken and whole exome sequencing (WES) and RNA-sequencing (RNA-Seq) performed. This allowed us to look for somatic mutations, copy number alterations and fusion genes. We also looked for the presence of mutational signatures, viruses, and putative germline predisposition alleles.

## MATERIALS AND METHODS

### Sample collection and nucleic acid isolation

The samples were FFPE tissues that had been collected as part of routine diagnostic procedures. Representative haematoxylin and eosin (H&E)-stained sections of the tissue blocks were independently reviewed by two specialist dermatopathologists (T.B. and I.F.) to confirm diagnoses and identify areas for sampling. Genomic DNA and RNA was extracted from the tumour samples and genomic DNA was extracted from the normal samples, using the AllPrep DNA/RNA FFPE Kit (Qiagen) according to the manufacturer’s instructions.

### Sequencing, read alignment and quality control

Preparation of DNA/RNA sequencing libraries, whole-exome capture using SureSelect Human All Exon V5 baits (Agilent) and paired-end sequenced using the NovaSeq6000 platform (Illumina) to generate 101 bp reads, were all performed as previously described.^23^ DNA sequencing reads were aligned to the GRCh38 reference genome using BWA-MEM (v0.7.17)^24^ and PCR duplicates were marked using the samtools (v1.14)^25^ markdup function. For tumours with matched normal DNA, tumour-normal sample concordance (as well as cross-individual contamination) was assessed using Conpair (v0.2).^26^

RNA sequencing reads were aligned to the GRCh38 reference genome using STAR (v2.5.0c15)^27^ and ENSEMBL (v103) gene annotations, and data quality was assessed by using RNA-SeqQC 2.^28^ Samples were excluded on the basis of quality issues, as previously described.^23^

### Identification and annotation of somatic variants

Somatic mutations were identified using cgpCaVEMan (v1.15.2)^29^ for single nucleotide variants (SNVs), SmartPhase (v1.2.1)^30^ for multi-nucleotide variants (MNVs) and cgpPindel (v.3.10.0)^31^ for insertions/deletions (indels). The details of the specific parameters, input files, and flagging rules that were used for each algorithm and downstream variant filtering are previously described.^23^ For tumour samples without matched normal tissue for comparison, variant calling with cgpCaVEMan and cgpPindel were run with *in silico* data in place of sequencing data from a normal sample. Additional downstream filtering to remove germline variants and artefacts is described in the **Supplementary Methods**.

The Ensembl (v103) Variant Effect Predictor (VEP)^32^ tool was used to predict the protein consequences of base changes and indels, and add custom annotations from the COSMIC (v97),^33^ gnomAD (v3.1.2)^34^ and dbSNP (v155)^35^ databases. Memorial Sloan Kettering Cancer Center’s Precision Oncology Knowledge Base, OncoKB^TM^, was used to define ‘cancer genes’ (https://www.oncokb.org/cancer-genes; data update: 24/10/2024) and predict pathogenicity of the somatic mutations.^36,37^ Lollipop plots of *PIK3CA* and *PIK3R1* were generated using MutationMapper in cBioPortal (https://www.cbioportal.org/mutation_mapper).

### Identification of somatic copy number alterations

Somatic copy number alterations (SCNAs) were identified using ASCAT (v3.1.2)^38^ and the results were reformatted as used as input files for GISTIC2 (v2.0.23)^39^ to identify significantly recurrent SCNAs. The threshold for the residual q-value for both broad and focal alterations was set at 0.10. Samples were excluded using previously described parameters.^23^

### Mutational signature analysis

COSMIC mutational signature identification (v3.3) was performed using SigProfilerExtractor (v1.1.21)^40^ run in exome mode, using GRCh38 as the reference and opportunity genome, as previously detailed.^23^

### Identification of significantly mutated genes

To identify driver genes, we used dNdScv (v0.0.1.0, git commit ID 0633182)^41^ and OncodriveFML^42^ which detect genes under positive selection in cancer. Analysis was performed as previously described for the dNdScv^23^ and OncodriveFML^43^ algorithms, and to identify significant genes, we considered those that had a q-value < 0.1.

### Mutual exclusivity and co-occurrence of mutated genes

DISCOVER (r_v0.9.4)^44^ was used to test for co-occurring and mutually exclusive mutation of genes, as previously described.^43^

### Identification and annotation of germline variants

Following the Genome Analysis Toolkit (GATK) Best Practices workflow^45^, PCR duplicates from aligned WES reads were marked using the Picard Tools (v2.25.4; https://broadinstitute.github.io/picard/) MarkDuplicates function and germline variants were identified from normal samples using GATK (v4.2.6.1) ^46^ for cohort calling of SNVs and short indels, as previously described.^43^ To identify variants present within known germline cancer predisposition genes, we looked for variants in genes used by England’s National Health Service (NHS) Cancer National Genomic Test directory v7.2 June-2023 to diagnose cancer predisposition in patients [https://www.england.nhs.uk/wp-content/uploads/2018/08/Cancer-national-genomic-test-directory-version-7.2-June-2023.xlsx]. Only variants with predicted moderate or high impact over the encoded protein were reported.

### Identification of fusion genes

Fusion gene identification was performed using STAR-Fusion v1.10.1, which used STAR (v2.78a)^47^ alongside the Trinity Cancer Transcriptome Analysis Toolkit (CTAT) genome reference library StarFv1.10 for GRCh38 GENCODEv37(ENSEMBLv103) gene annotations. To assess the coding effect of the fusions, annotate their presence in previous databases and validate them *in silico*, we used FusionInspector v2.6.0 as previously described.^23^ Gene fusion predictions were excluded on the basis of previously published parameters.^23^

### Viral analysis

We used Kraken2 v2.1.2^48^ for pathogen identification in the tumour unfiltered RNA sequencing reads and unmapped DNA reads, as previously described.^43^ Our cut off for significance was P <0.05 and log10 clade-levels reads >0.1.

## RESULTS

### Demographics and phenotype of the HP cohort

The HP cohort consisted of 68 FFPE primary tumour samples from 68 patients (all females; with a mean ± SD age of 46 ± 11 years), ascertained from 8 institutions across 7 countries. All the tumours were located in the anogenital region, with the most common site being the vulva (including tumours classified as coming from the clitoris and labia; 59/68, 87%), followed by the anus/perianal area (4/68, 6%; **Table S1**; see Supporting Information). Histologically, HP presented as a well demarcated nodular tumour in the dermis, showing a protrusion of papillary structures into cystic spaces (**Figure S1**; see Supporting Information). The papillae contained a fibrous core and were lined by a double cell layer composed of inner small myoepithelial cells and outer columnar cells, focally exhibiting decapitation secretion or oxyphilic metaplasia. Rare mitotic figures were observed. In some cases, the overlying epidermis was acanthotic, and rarely ulcerated. Connection to the underlying tumour may be an additional finding. An admixed chronic inflammatory infiltrate was observed in a subset of cases.

The HP cohort was subdivided into cases that had tumour tissue with matched normal tissue (peri-lesional tissue or tissue from another site; n=35), which represented our ‘discovery’ cohort, and cases that had tumour tissue only (i.e., no matched normal was available for analysis; n=29), which represented our ‘validation’ cohort. Whole-exome sequencing was performed on the DNA from both groups. Pulldown RNA-sequencing was performed on tumour RNA from 60 cases (cases were from both the discovery and validation cohorts). Taken together, this allowed us to look for somatic mutations, copy number alterations and fusion genes. We also looked for the presence of mutational signatures, viruses, and putative germline predisposition genes. A summary of sample details, including clinical information, as well as which samples were used for each of the analyses, is provided in **Table S1** (see Supporting Information).

### *PIK3CA* and *PIK3R1* are mutually exclusive HP driver genes

Focussing on the discovery cohort, the tumour mutational burden (TMB) of HP was very low (median: 0.270 mutations per megabase, range: 0.083 – 0.498 mutations per megabase). As such we were not able to identify any COSMIC mutational signatures. The most recurrently mutated genes were *PIK3CA* (27/35, 77% cases) and *PIK3R1* (6/35, 17% cases; **Figure 1**). *PIK3CA* and *PIK3R1*, respectively, encode the p110α catalytic subunit and p85α regulatory subunit of the phosphatidylinositol 3-kinase (PI3K) heterodimer. The predominance of *PIK3CA* and *PIK3R1* mutations were mirrored in the validation cohort (Figure S2). Considering both cohorts, the *PIK3CA* mutations were predominantly missense mutations, all located at known hotspots^49^ (p.H1047R/L and p.E545K), and are well-characterised activating/oncogenic mutations (**Figure 2a** and **Table S2**; see Supporting Information). Similarly, all but one of the mutations observed in *PIK3R1* occurred at known hotspots within the iSH2 domain (one of three key domains that inhibit the catalytic subunit of PI3K) and were predicted by OncoKB^TM^ to be ‘likely oncogenic’ in function (**Figure 2b** and **Table S2**; see Supporting Information).

**Figure 1.**
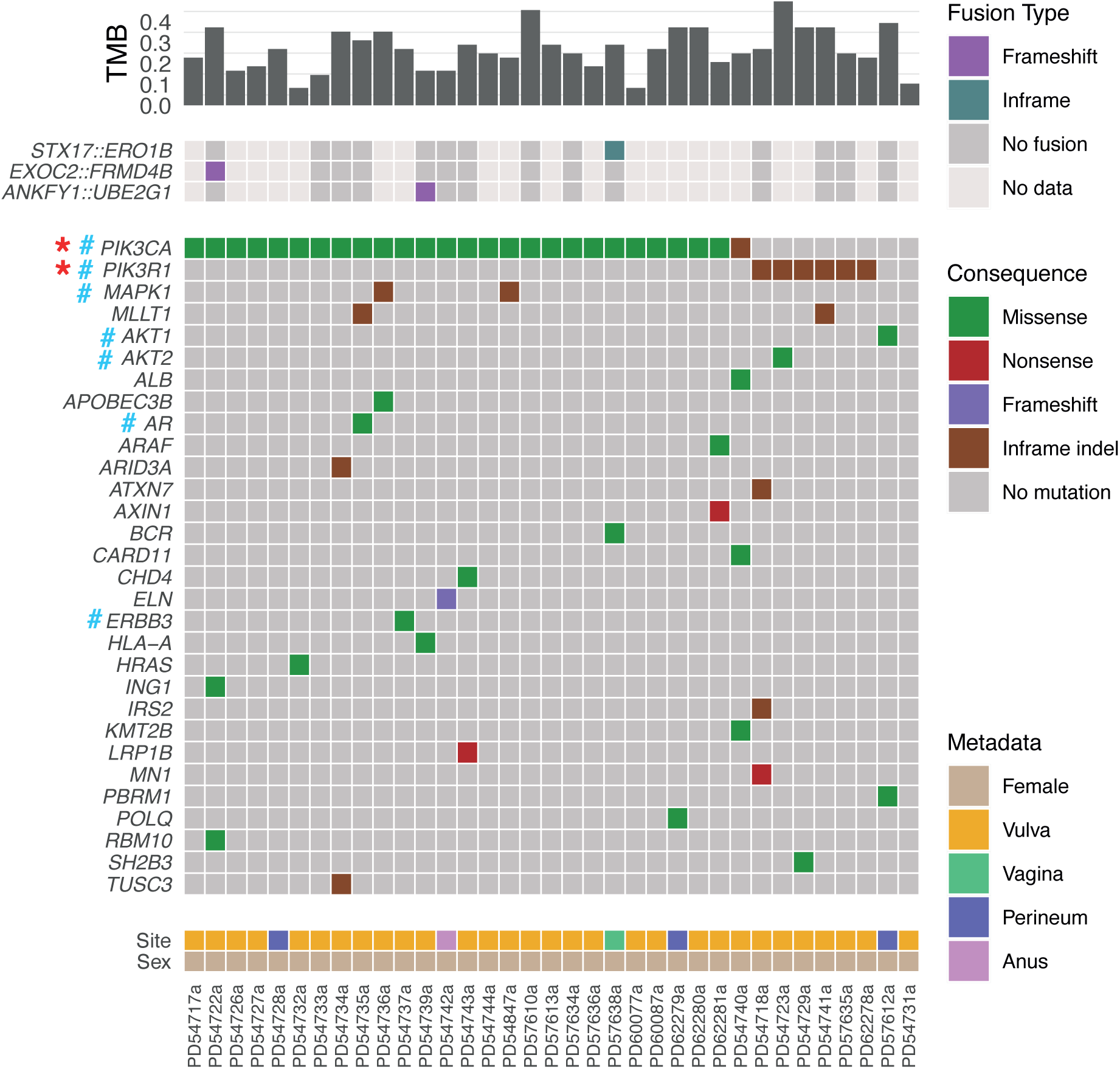
Overall mutational landscape of hidradenoma papilliferum. The ‘discovery cohort’ (n=35). Tumour mutational burden (TMB) is shown as mutations per megabase. The oncoplot panel shows cancer genes in the OncoKB database for which a mutation was present in at least one sample. Asterisks (in red) indicate significantly mutated genes and hashtags (in light blue) indicate genes in the PI3K/AKT/mTOR signalling pathway.

**Figure 2.**
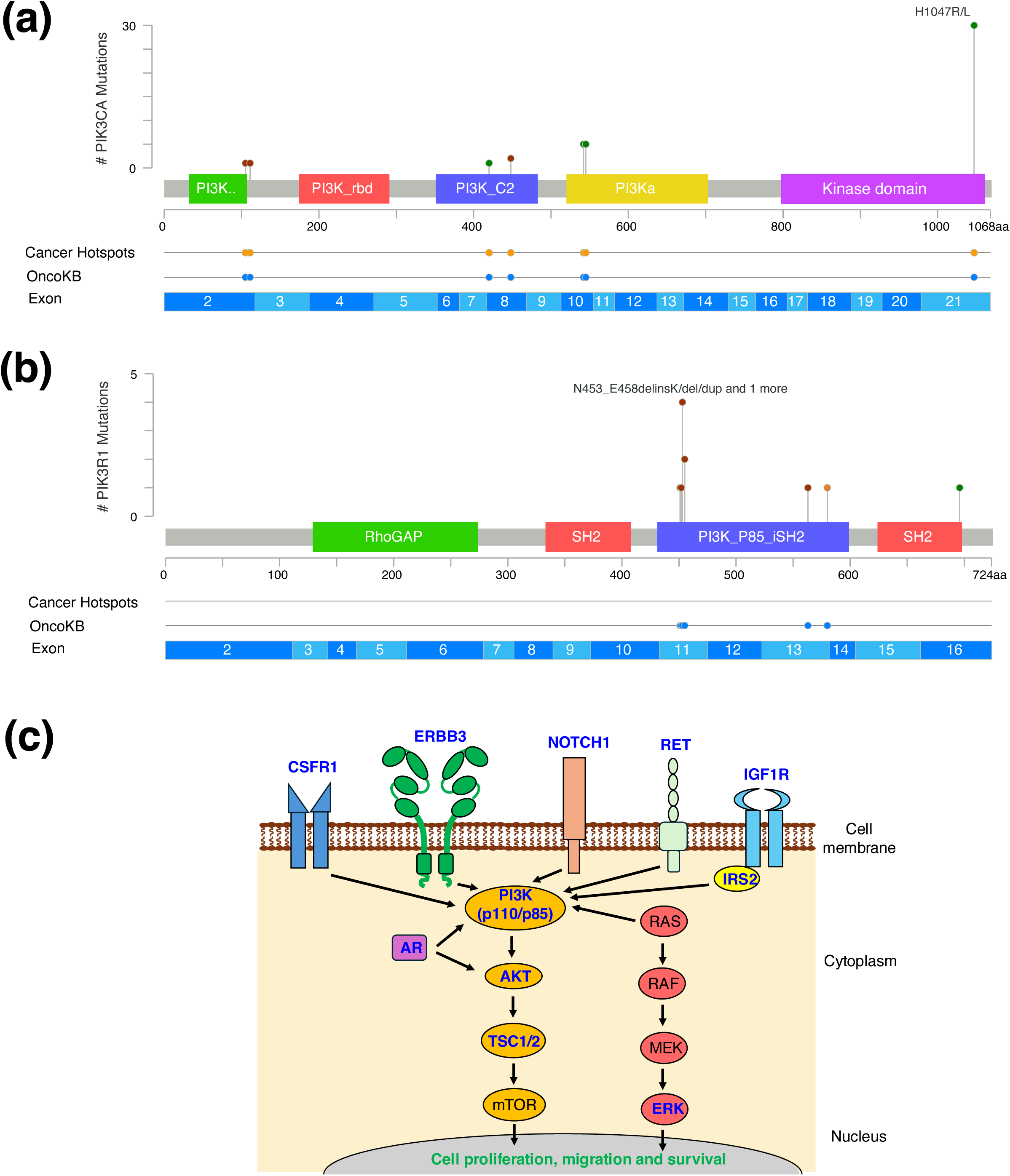
Importance of the PI3K/AKT/mTOR signalling pathway in hidradenoma papilliferum (HP). Lollipop plots showing the location of mutations in **(a)** *PIK3CA* and **(b)** *PIK3R1* in the discovery and validation HP cohorts. Shown are the protein domains (colour boxes) and the exons (numbered blue boxes). Also indicated is if the mutation is a known ‘hotspot’ in cancer and whether the mutation is present in the OncoKB cancer database. **(c)** Schematic diagram showing the PI3K/AKT/mTOR signalling pathway and associated pathways and interactors. Blue font indicates the translated proteins of genes that were altered (somatic mutations or germline variants) in the HP cohort. *MAPK1* encodes ERK2 (shown as “ERK”).

Using the discovery cohort, two different algorithms were employed to identify genes under positive selection in the tumours. *PIK3CA* and *PIK3R1* were found to be significantly mutated (q = 0 and 1.1 x 10^-12^, respectively, for dNdScv, and 1.6 x 10^-5^ and 4.6 x 10^-3^, respectively, for OncodriveFML), and thus represent candidate driver genes of HP. Co-mutation and mutual exclusivity analysis found that mutations in *PIK3CA* and *PIK3R1* were mutually exclusive (q = 5.1 x 10^-3^).

Other recurrently mutated genes included *MAPK1* and *MLLT1* (2/35, 6% cases; **Figure 1**) in the discovery cohort, and *ZFHX3* and *RECQL4* (**Figure S2**; see Supporting Information) in the validation cohort.

### Involvement of the PI3K/AKT/mTOR signalling pathway in HP

The PI3K enzyme is a key part of the PI3K/AKT/mTOR intracellular signalling pathway responsible for driving cell proliferation, migration and survival (**Figure 2c**). A previous study suggested that tumourigenic alterations in the PI3K/AKT/mTOR-pathway were indispensable in HP, with cases either having hotspot mutations in *PIK3CA* or *AKT1* (n=5 cases), or mutations in at least one gene contributing to or involved in PI3K/AKT signalling (n=7 cases).^14^ Considering both our discovery and validation cohorts, we identified recurrent mutations in key members of this pathway, specifically *PIK3CA*, *PIK31R* and *AKT1*. In agreement with a previous study, we observed that *PIK3CA* and *AKT1* mutations were not found in the same tumours^20^, similar to that seen with *PIK3CA* and *AKT2* mutations (**Figure 1**). However, mutual exclusivity could not be statistically assessed with only a single tumour showing either an *AKT1* or *AKT2* mutation in our discovery cohort. Other known cancer genes with a link to the PI3K/AKT/mTOR signalling pathway that were mutated in our discovery cohort were either receptors or receptor-associated proteins that signal via the PI3K/ATK/mTOR pathway (*ERBB3, IRS2*), or are members of pathways that cross-talk with the PI3K/AKT/mTOR pathway (*AR* and *MAPK1*; **Figure 2c** and **Table S3a**; see Supporting Information). Of these four genes, only *AR* has been previously reported as mutated in HP.^14^

### Copy number alterations are not driver events in HP

To-date, no SCNA investigations have been performed in HP. Using the discovery cohort, we found the SCNA landscape of HP was very quiet, with no recurrent chromosomal gains, losses or copy neutral loss of heterozygosity (cnLOH; **Figure S3**; see Supporting Information). The ploidy estimates of the tumours were all diploid. Taken together, this strongly suggests that SCNAs are not driver events in HP.

### Identification of fusion genes found in HP

Fusion genes are an important class of driver event in cancer, as they can play a role in both tumour initiation and progression. Using data from the RNA sequenced samples (n=60), a total of five different fusion genes were identified in our HP cohort. These included two frameshift fusions (*EXOC2::FRMD4B*, *ANKFY1::UBE2G1*) and three in-frame fusion (*IQGAP1::ZNF774*, *SMARCD3::ASXL2*, *STX17::ERO1B*; **Figure 1** and **Table S4a**; see Supporting Information). *EXOC2::FRMD4B* and *IQGAP1::ZNF774* have been previously been identified in other tumour types,^50,51^ whilst *ANKFY1::UBE2G1, SMARCD3::ASXL2* and *STX17::ERO1B* have not previously been reported, although each of these genes have been found as fusion gene partners in a range of tumour types.^52^ Given these fusions were found in tumours with somatic *PIK3CA* or *AKT1* mutations, and the fusions were only found in one tumour sample each, it would suggest they are not prominent driver events.

### Germline susceptibility in HP

As we sequenced matched normal tissue for tumours in our discovery cohort, we were able to search for putative pathogenic germline variants (single nucleotide variants and insertions/deletions) in HP (**Table S5**; see Supporting Information). To focus on clinically relevant genes, we considered only known cancer-predisposition genes with variants of moderate or severe functional impact as estimated by VEP (see **Methods**). Using these criteria, the most altered genes were *ARID1B* and *CEBPA* (3/35, 8.6% cases each), with in-frame deletions (**Figure S4**; see Supporting Information). Genes found altered in two patients included *ATM, CDKN1A, MET, PMS2* and *POLE*). Importantly, there was a patient carrying a ClinVar ‘pathogenic’ variant, specifically PD54740 (a frameshift deletion in *NF1* at p.L1877X; **Figure S4**; see Supporting Information). There were also patients with variants in members of the PI3K/AKT/mTOR signalling pathway (*TSC1/2*), variants in receptors that signal through the PI3K/AKT/mTOR pathway (*CSFR1, NOTCH1, RET, IGF1R*) or variants in members of pathways that cross-talk with the PI3K/AKT/mTOR pathway (*MAPK1*; **Figure 2c** and **Table S3b**; see Supporting Information).

### No significant evidence of HPV in HP

As HPV infections are common and the presence of HPV DNA can readily be found in anogenital epithelial samples, it is not surprising that HPV has been reported in HP samples.^11,12^ As a potential role for HPV in HP pathogenesis remains unclear, we looked for the presence of HPV, or any other viruses, in the DNA and RNA sequencing reads of the HP samples. Using the Kraken2.0 algorithm which searches for short segments of viral sequences (‘minimisers’), we did not find any significant levels of minimisers belonging to the HPV genera (*alphapapillomavirus, betapapillomavirus*, or *gammapapillomavirus*).

## DISCUSSION

PI3Ks are a family of enzymes, composed of for four different classes. Class I PI3Ks are heterodimers composed of a p110 catalytic and p85 regulatory subunit, and are involved in cellular functions such as growth, proliferation, differentiation, migration and survival. Many of these cellular functions relate to the ability of the class I PI3Ks ability to activate AKT, i.e., the PI2K/AKT/mTOR signalling pathway. Members of this pathway are frequently mutated in numerous cancer types.^53^ It has previously been suggested that the PI3K/AKT signalling pathway is important in HP tumourigenesis.^14^ In our study, the majority of samples had mutations in PI3K/AKT/mTOR family members, with 34/35 (97%) of our discovery cohort tumours having somatic mutations in either *PIK3CA, PIK3R1, AKT1* or *AKT2*. Furthermore, due to our large cohort size, we were able to perform statistical analyses and found that mutawon of *PIK3CA* and *PIK3R1* were under posiwve selecwon, thus, implicawng these as driver genes of HP. Moreover, mutations in these genes were statistically identified as being mutually exclusive.

*PIK3CA* mutations can have clinical significance, thus, it is important to note that the mutations in *PIK3CA* observed in the HP cohort all occurred at hotspots that are known to activate the protein, resulting in oncogenic activity. Similarly, all but one of the mutations observed in *PIK3R1* in our HP cohort occurred at known hotspots and were predicted to be ‘likely oncogenic’ in function. We also identified recurrent somatic mutations in *ERBB3* (ERBB3 contains multiple binding sites for p85α, thus allowing direct activation of the PI3K signalling pathway). In addition, there were somatic or germline mutations in PI3K/AKT/mTOR signalling pathway members (*TSC1/2*), receptors or receptor-associated proteins that signal through the PI3K/AKT/mTOR pathway (*IRS2*, *CSFR1, NOTCH1, RET* and *IGF1R*), or members of pathways that crosstalk with the PI3K/AKT/mTOR pathway (*AR* and *MAPK1*).

Previous studies have reported individual HPs with a mutation in a variety of cancer genes, including *BRAF* (p.V600E),^15,20^ *APC,*^15^ *ERBB4,*^15^ and *KMT2C,* ^14^ among numerous others, however, we did not find mutations in any of these genes in our cohort. Although many of these studies screened the *RAS* genes for hotspot mutations, none were identified,^15,20,21^ however, we found a sample with a *HRAS* mutation (at hotspot p.G13V), as well as a hotspot *PIK3CA* mutation (p. H1047R; PD54732a). Yet, *HRAS* playing a key role in driving HP seems unlikely with only a single sample showing a mutation.

In many tumour types, fusion genes can represent driver events. We identified five different fusion genes in our HP cohort, two of which have been previously identified in large scale fusion gene screens; *EXOC2::FRMD4B* in a squamous cell carcinoma sample^50^ and *IQGAP1::ZNF774* in a thyroid adenocarcinoma sample.^51^ However, since the five fusion genes were only found in a single tumour each, and all occurred in tumours with somatic mutations in the PI3K/AKT signalling pathway, it is unlikely they represent driver events in HP.

The aetiology of HP is currently unclear, and reports of HPV DNA in some anogenital HP lesions^11,12^ has led to the suggestion that HPV may play a causative role. However, we found no evidence of HPV, or any viral sequences in either our WES or RNA sequencing datasets. Thus, in agreement with other studies which also did not find any HPV DNA in their HP samples,^13–15^ it suggests that HPV does not play a causative role in driving HP.

It is interesting to note that many of the previous HP sequencing studies had cases for which no mutations could be identified, due to the use of hotspot Sanger sequencing or targeted cancer panel sequencing only.^15,20–22^ For example, mutations in *ERBB3* have not been previously reported in HP as *ERBB3* was not included in the targeted panels employed in these studies.^14,15,22^ This underscores the importance of WES to obtain a comprehensive mutational landscape. Nevertheless, although the exome is estimated to contain ∼85% of mutations with large effects on disease-related traits,^54^ a limitation of WES over whole genome sequencing (WGS) is that we will not have uncovered mutations in introns, promoters and other regulatory regions that may play a role in the development of HP. Similarly, WES (and pulldown RNA sequencing) could have limited our ability to capture viral sequences that may have been present in some samples. Finally, future molecular profiling studies of HP would benefit from including the rare cases of HP from male patients (to be inclusive) and HP arising in non-anogenital areas (as a comparator).

Our rich understanding of the pathophysiology of cutaneous tumours has occurred in large part through molecular profiling of tissue samples to identify key somatic driver events, candidate germline pre-disposition genes/alleles and provide insight into potential aetiological factors (such as mutagenic exposures and viruses). Our comprehensive characterisation of the genomic landscape of a large cohort of HP has identified *PIK3CA* and *PIK3R1* as mutually exclusive driver genes and confirmed a key role for the alteration of genes in the PI3K/AKT signalling pathway in driving this tumour type.

## Acknowledgements

The authors thank DNA Pipelines at the Wellcome Sanger Institute for preparation of the libraries, pulldown and sequencing of the DNA/RNA samples.

## Data availability

The sequencing data is available in the European Genome and Phenome Archive (EGA) repository (https://www.ebi.ac.uk/ena/browser/home) under the study accessions EGAD00001015481 for whole exome data, and EGAD00001015480 for RNA-seq data.

## Ethics statement

Ethical approval for the use of all patient samples in this project was obtained by a local committee at the institution of origin and from Research Governance at the Wellcome Sanger Institute. This study is part of the DERMATLAS Project that has been approved by the NHS Health Research Authority; Research Ethics Committee (REC) reference: 21/PR/1024, IRAS project ID: 304621. The study is anonymised linked and patient consent was given for inclusion of their tissue samples and associated clinical metadata in the study.

## SUPPORTING INFORMATION

**Figure S1.**
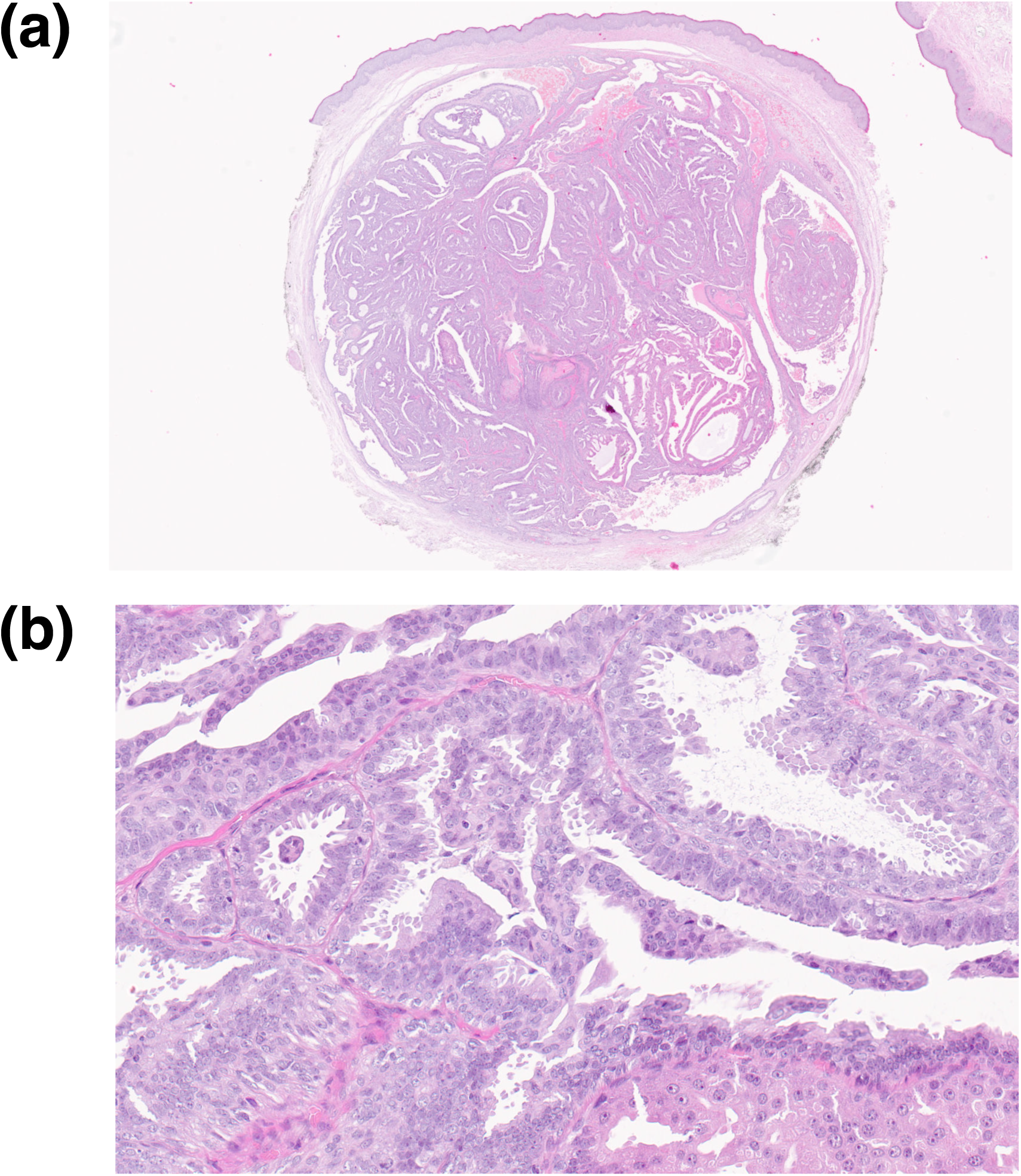
Histological presentation of hidradenoma papilliferum. Representative haematoxylin and eosin-stained sections showing a **(a)** well-circumscribed dermal nodule (1.25x magnification), **(b)** characterised by papillae projecting into cystic spaces and covered by a double cell layer including inner small myoepithelial cells and outer columnar cells showing decapitation secretion or oxyphilic metaplasia (20x magnification).

**Figure S2.**
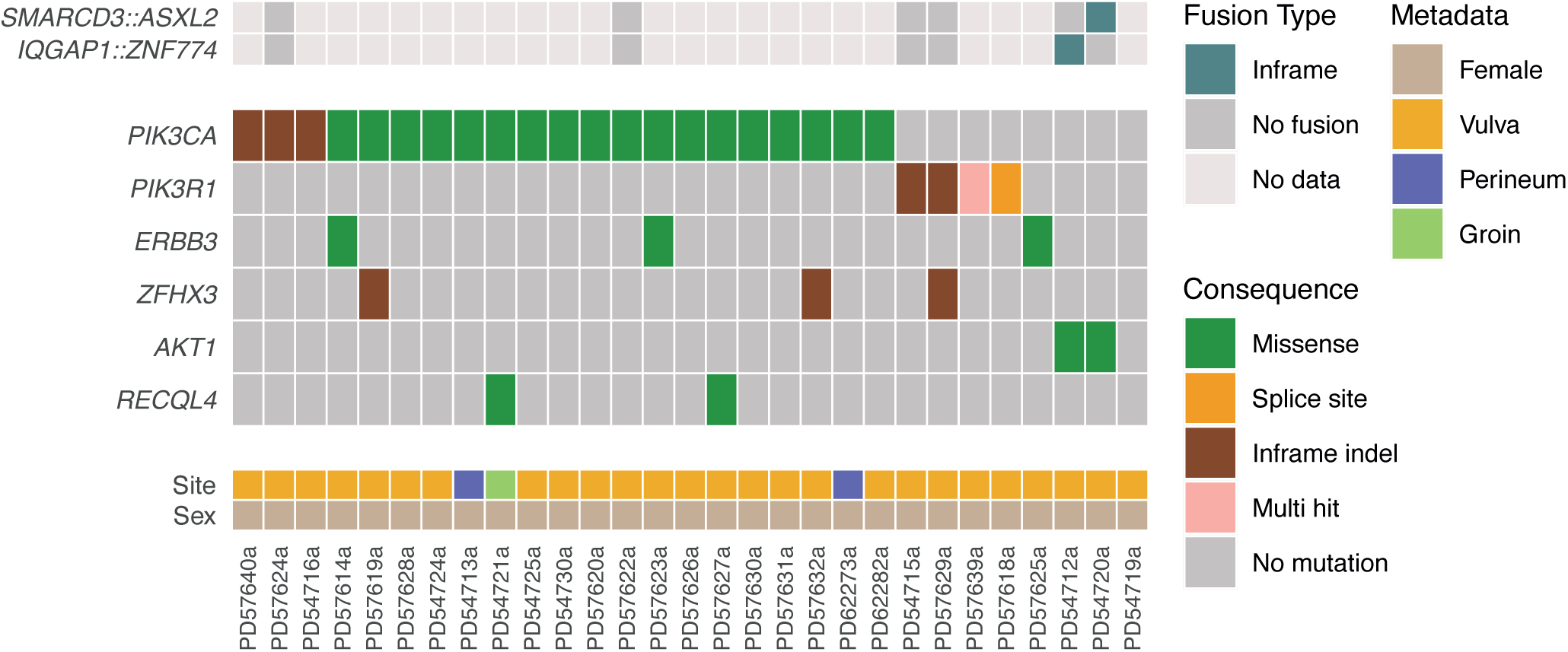
Overall mutational landscape of hidradenoma papilliferum. The oncoplot panel shows OncoKB cancer genes for which a mutation was present in at least two samples in the validation cohort (n=29).

**Figure S3.**
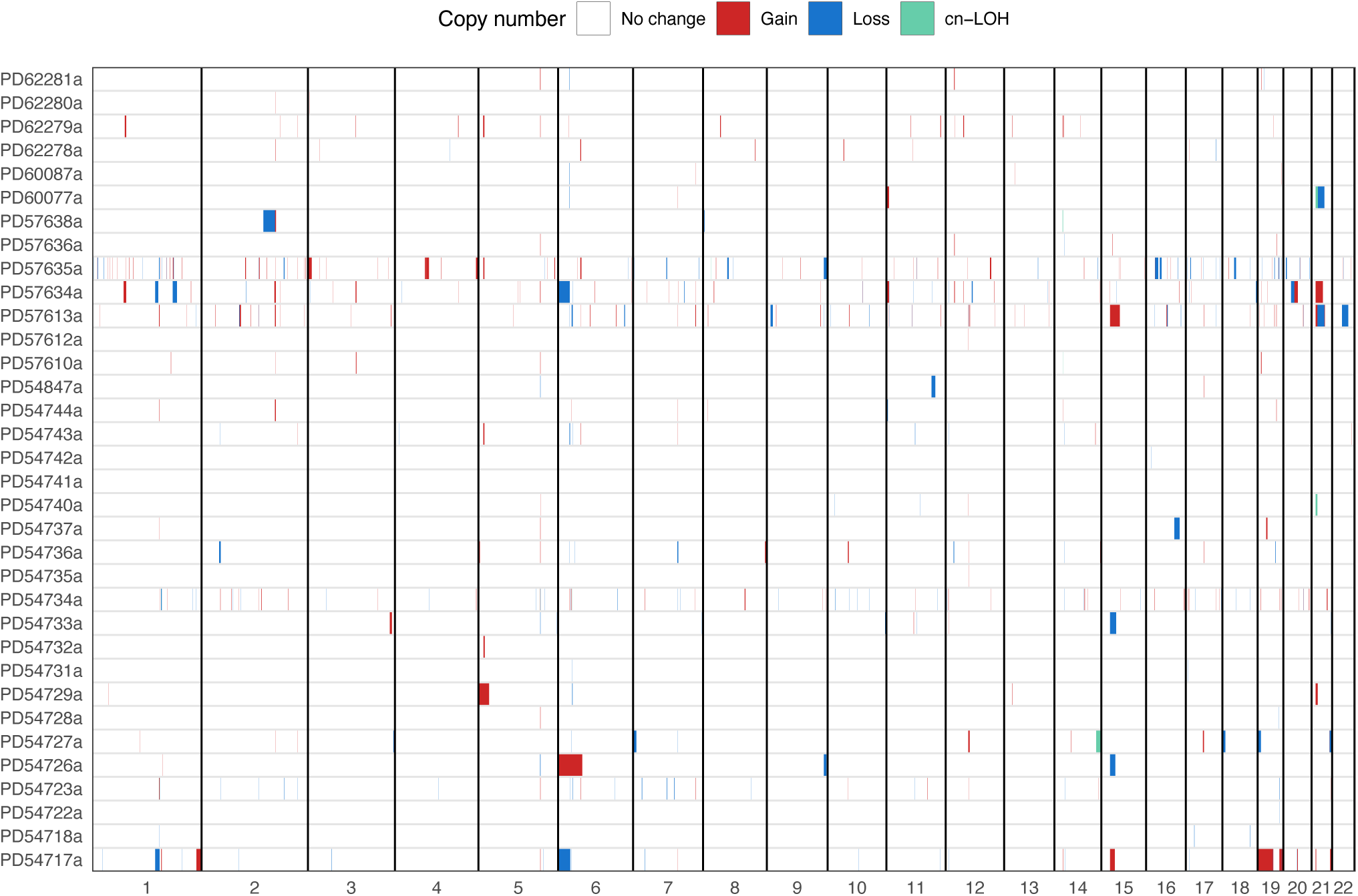
Somatic copy number alterations in hidradenoma papilliferum. Plot showing the frequency of somatic copy number alterations in HP (n=34, discovery cohort), using 1 megabase windows.

**Figure S4.**
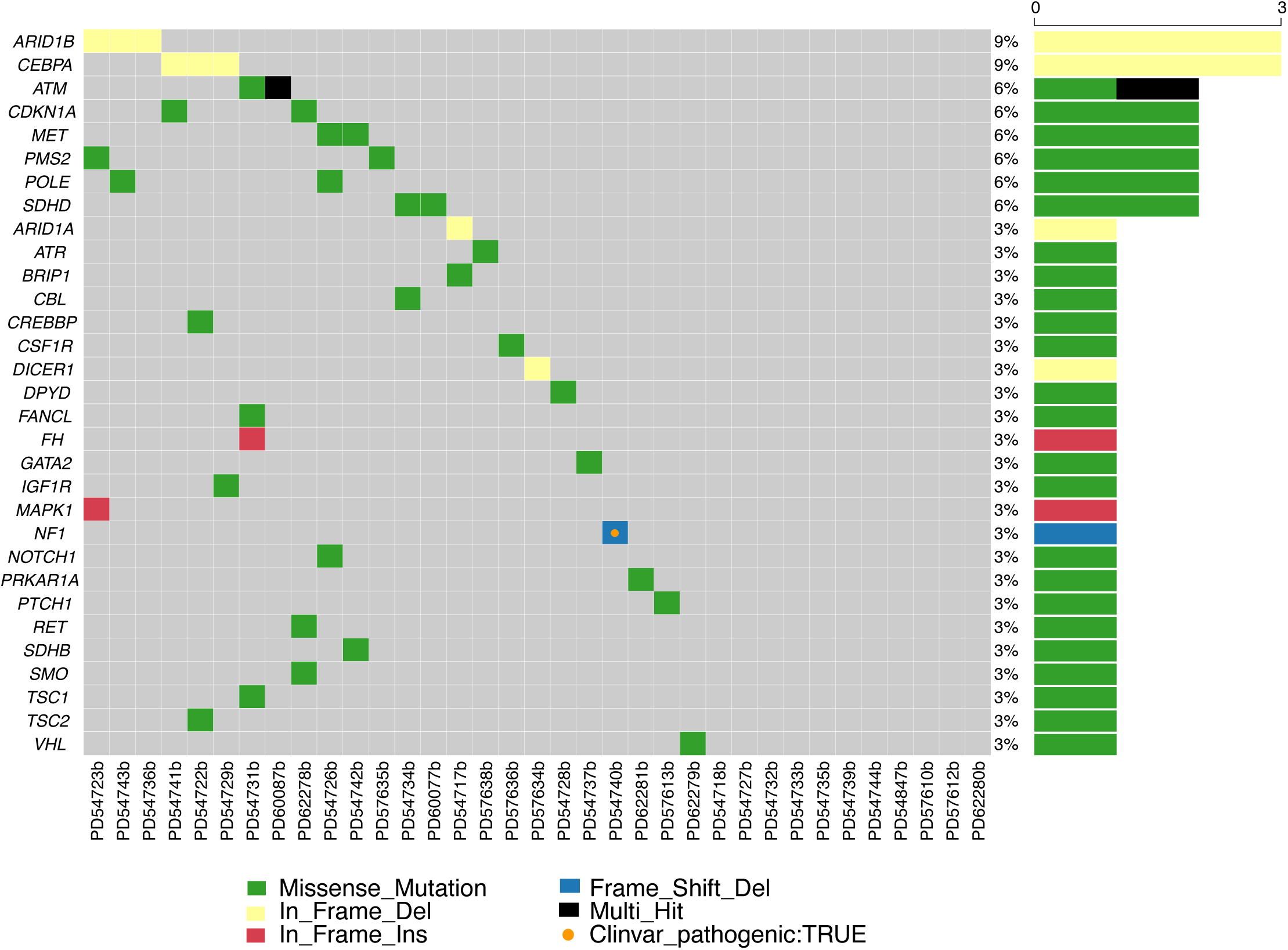
Germline alterations of cancer-predisposition genes in hidradenoma papilliferum patients. Oncoplot showing germline variants with predicted moderate or high functional impact in the hidradenoma papilliferum samples (n=35) affecting a gene used by National Health Service (NHS) England for diagnosis of cancer predisposition through genetic/genomic testing. “Multi_Hit” indicates the presence of more than one mutation in the gene for that sample and mutations reported as “Pathogenic”, “likely_pathogenic” or “risk_factor” in the ClinVar database are marked with an orange circle.

**Table S1. Details of the hidradenoma papilliferum samples used in the study.** Sex is male (M) or female (F). Age is age at diagnosis (in years). Site_specific is the location of the tumour on the body (L, left; R, right). Site_group is head and neck, trunk or extremities. Abbreviations: indel, insertion/deletion; MNV, multi-nucleotide variant; SNV, single nucleotide variant.

**Table S2. Somatic mutations in hidradenoma papilliferum.** Lists containing the set of somatic short indels, single and multi-nucleotide mutations identified in the hidradenoma papilliferum samples from the (a) ‘discovery’ cohort (n=35), and (b) ‘validation’ cohort (n=29). Variant information on this table follows the column format specified for the Mutation Annotation Format (MAF) v1.0.0 and Variant Call Format (VCF) v4.1, with additional consequence and functional prediction information obtained from Variant Effect Predictor (VEP) annotations. Additional information such as: SIFT and POLYPHEN scores and categories, domain information, and previous information for the variant from dbSNP, COSMIC, ClinVar and gnomAD are stated. Tumour_ID/Normal_ID indicate sample IDs of matched tumour/normal samples. For each mutation, the VEP predicated variant consequence relative to the gene’s canonical transcript is shown. In instances where multiple consequences were identified Main_consequence_VEP shows the single most deleterious consequence.

**Table S3. Somatic mutations and germline variants in PI3K/AKT/mTOR pathway-associated members in hidradenoma papilliferum. (a)** Somatic mutations and **(b)** germline variants identified in PI3K/AKT/mTOR signalling pathway members, receptors that signal through the PI3K/AKT/mTOR pathway, genes that regulate the PI3K/AKT/mTOR pathway or genes in pathways that cross-talk with the PI3K/AKT/mTOR pathway.

**Table S4. Gene fusions in hidradenoma papilliferum.** Summary of all the gene fusions identified in the hidradenoma papilliferum samples (n=60). All fusions reported have been annotated and passed the filtering criteria. Gene symbols are supplied, along with Ensembl gene IDs.

**Table S5. Germline mutations in hidradenoma papilliferum.** Table containing the summary of germline mutations within the normal tissue of the hidradenoma papilliferum samples (n=35). Only mutations that passed our variant quality filtering criteria, had moderate or high predicted protein consequences, and were located within genes previously associated with cancer predisposition are reported.

## Supplementary methods

### Filtering of variants from unmatched tumour samples

For variant calls from unmatched tumour samples (those with out a matched normal sample), we leveraged population databases and the unfiltered variant calls from unmatched and matched tumour samples to remove probable common SNPs and artefacts. In addition to the filtering using dbSNP and gnomAD variants described for variants in matched tumour samples, a variant was classified as a germline variant or an artefact if it met any of the following criteria: (1) Low-quality variants (see below). (2) Recurrent artefacts. The variant is present in unmatched tumour and all have a variant allele frequency (VAF) < 0.1, or, VAF < 0.1 and >50% of unmatched cases and tumour coverage < 35x in two thirds of cases. (3) Germline variants. In normal samples, the variant is in 4 or more samples (making up >= 5% of samples), or, if in <= 3 normal samples and the variant is flagged as low-quality. (4) Alignment artefact. an SNV overlaps a recurrent germline indel by < 5 bp, or, and indel overlaps a recurrent germline indel of any size. (5) Low-quality multi-nucleotide variant (MNV) (see below). (6) Variant consequence. The variant does not alter the protein sequence or splice site, or does result in a retained stop or start codon. The variant is rescued, however, if it is present in the Cancer Hotspot database. In order to identify the highest confidence somatic variants, in addition to variants that were known cancer hotspot mutations and removing germline variants and artefacts, we retained variants that met at least one of the following criteria: (1) the variant affects an OncoKB cancer gene; (2) the variant is not in either the dbSNP or gnomAD databases; or, the population AF is not >= 0.001 in both dbSNP and gnomAD; or, the variant is not flagged in gnomAD as low-quality; (3) the variant is in the ClinVar database and the clinical significance is ‘likely pathogenic’ or ‘pathogenic’; or, the SIFT and PolyPhen predictions are ‘deleterious’ and ‘probably damaging’, respectively.

In some cases, artefacts and germline SNPs may be called as high-quality variants in some samples, and low-quality in others. We used unfiltered variant calls from all matched and unmatched tumours to identify variants in the unmatched tumours that may have been flagged as low-quality in other samples. If any of the following condtions were met, then the variant was removed as a germline variant or artefact. (1) If, in all matched tumour samples with the variant, the variant is flagged as low-quality, then the variant is removed. (2) If, in the unmatched tumour samples, the variant is seen in 3 or 4 samples and is flagged as low-quality is all of them, then the variant is removed. (3) If, in either the matched or unmatched tumour samples, the variant is seen in 5 or more samples and flagged as low-quality in at least 25% of samples, then the variant is removed. For multi-nucleotide variants that were called by merging of phased, adjacent SNVs using casmsmartphase (v0.1.8), if any of the SNVs constituting the MNV was flagged originally by cgpCaVEMan (v1.15.2) as low-quality, then the MNV is flagged as low-quality.

